# Hydroxychloroquine treatment for recurrent pregnancy loss: a study protocol for a randomized, double-blind, placebo-controlled trial

**DOI:** 10.1101/2024.05.23.24307818

**Authors:** Anna L. Talbot, Pia Egerup, Anne L. Lunøe, Karen K. Jensen, Marie L.T. Chonovitsch, Anne-Sofie Korsholm, Ida Behrendt-Møller, Tanja S. Hartwig, David Westergaard, Kilian Vomstein, Elisabeth C. Larsen, Barbara M. Fischer, Henriette S. Nielsen

## Abstract

**Introduction:** Pregnancy loss occurs in 25% of all pregnancies. While 50-60% of the pregnancy losses are due to fetal chromosomal and structural abnormalities, incompatible with life, some of the remaining euploid pregnancy losses are likely caused by maternal conditions. At least 2% of all women suffer ≥3 consecutive pregnancy losses, defined as recurrent pregnancy loss (RPL). A hyperactive maternal immune system has been suspected to be responsible for rejection of healthy fetuses and be the underlying cause for unexplained recurrent pregnancy loss. However, very sparsely evidence-based treatment is available. The anti-inflammatory immune modulatory effects of hydroxychloroquine (HCQ) and casuistic reports of live births by women with previous RPL after HCQ-treatment, have given rise to several hypothesizes for HCQs impact on RPL. In this multicenter randomized, double-blinded, placebo-controlled trial (db-RCT) we aim to uncover whether HCQ can increase the live birth rate (LBR) in women with RPL, thus laying the foundation for an evidence-based treatment to make a real difference for a much-overlooked group of patients.

**Methods and analysis:** We will assess HCQ treatment in women with a minimum of four consecutive unexplained pregnancy losses (or three pregnancy losses including a second trimester pregnancy loss), and monitor a potential difference in LBR among women allocated treatment with HCQ and placebo, without exclusion of any included patients (Intention to treat analysis) and with exclusion of those with ectopic pregnancy, pregnancy loss due to chromosome abnormalities, neglect of treatment, induced abortion or those who withdraw from the treatment earlier than the protocol dictates (per protocol analysis). Secondary outcome measures include comparison of the rate of perinatal complications in the intervention group to the placebo group and examination of inflammatory and immunological mechanisms in peripheral blood. We aim to include 186 patients in the db-RCT. In addition, we will in a sub-study with 10 women monitor metabolic activity measured by ultra-low dose FDG-PET/CT scans before and during pregnancy. Metabolic activity correlates with grade of inflammation and will be measured in the uterus and immune-related tissue such as bone marrow, spleen, thymus, and lymph nodes.

**Ethics and Dissemination:** The db-RCT including the PET/CT sub-study has been approved by The Regional Committee on Health Research Ethics, The Danish Medicines Agency, The Danish Data Protection Agency, and transferred to The Center for Data handling Capital Region of Denmark in May 2024. The trial conforms to good clinical practice guidelines (GCP) and has been registered at ClinicalTrials.gov (ClinicalTrials.gov ID NCT03305263). Findings will be disseminated through peer-reviewed journals and at professional conferences.

**Strengths and limitations of this study:**

- Assessment of HCQ treatment in women with RPL to potentially increase LBR.
- Assessment of the rate of perinatal outcomes associability with HCQ treatment.
- RPL trials so far have struggled to obtain pregnancy tissue from PL and if obtained been severely contaminated with maternal tissue. We will use circulating cell-free fetal DNA (cffDNA), isolated from maternal peripheral blood for fetal genetic diagnostics as we demonstrated feasible in our recent Lancet publication.
- Non-invasive monitoring of inflammation in the uterus before and during pregnancy by the means of ultra-low dose PET/CT scans.

## INTRODUCTION

Pregnancy loss (PL) occurs in 25% of all pregnancies. A PL is defined as the spontaneous demise of a pregnancy prior to the fetus reaching viability, and the majority occurs within the first twelve gestational weeks^1^. While 50-60% of the PLs are due to fetal chromosomal and structural abnormalities, rendering the fetuses unable to live, some of the remaining euploid PLs are likely caused by maternal conditions^2^. At least 2% of all women suffer ≥3 consecutive PL, which is defined as recurrent PL (RPL), albeit the definition varies by country. The frequency of fetal chromosomal abnormalities decreases with increasing number of PL^3^, resulting in a higher risk for maternal cause to the PL for women with PRL. Less than half of RPL-cases can be explained by uterine abnormalities, parental balanced chromosomal rearrangement or coagulatory, endocrine, or autoimmune conditions^1^. The remaining couples are diagnosed with unexplained RPL. Based on accumulating evidence, an abnormal immune reaction from the mother towards the fetus can be the underlying mechanism of pregnancy rejection in some of the unexplained RPL cases^1,4–10^. This has fostered the idea of immunomodulatory therapies such as prednisolone, intravenous immunoglobulin-treatment, lymphocyte immunization, and intravenous lipid-infusion to increase live birth rate (LBR)^5,11,12^. Unfortunately, many of the studies are uncontrolled or deliver controversial results, and as a result there is very sparse clinical evidence for the effects of these treatments for unexplained RPL.

Hydroxychloroquine (HCQ) is an antimalaria and anti-inflammatory drug originally used in the treatment and prevention of malaria. Based on the anti-inflammatory immune modulatory effects. HCQ has extended indications to also treat autoimmune conditions. HCQ is registered for treatment of rheumatoid arthritis, Lupus erythematosus disseminates, polymorph rash from sun exposure, and juvenile idiopathic arthritis. HCQ is less toxic than Chloroquine and has only few side effects, with HCQ-Induced Retinal Toxicity being the most serious^13^. Some studies including more than 10.000 pregnant woman have among other treatments investigated the eligibility of Chloroquine treatment for pregnant women as malaria prophylaxis and have not found an increased risk for unwanted fetal effects^14,15^. Data on more than 800 pregnant women treated with HCQ for immune modulation showed no increase in unwanted effects on the fetus/offspring^13^.

The anti-inflammatory immune modulatory effects of HCQ^16,17^ and casuistic reports of live births by women with previous RPL after HCQ-treatment^18,19^, have given rise to hypothesizes of several mechanisms for how HCQ can reduce the risk for PL in RPL^15^. HCQ is suspected to prevent systemic inflammation, oxidative stress, endothelial dysfunction, and hypercoagulability in pregnant women-states presumed associated with RPL^15^. The potential immune mechanisms of HCQ in preventing RPL include I) inhibition of Toll like receptors (TRL)^15,17,20^, altering the T-cell activation and proliferation and consequently decreasing the levels of circulating TNF-α^15,21,22^, interferon-gamma^15,22^, and different interleukins^15,22^ promoting the switch to a TH2 phenotype of a healthy pregnancy ^15^, and II) by inhibiting platelet aggregation reducing the formation of blood clots^15^.

At The Recurrent Pregnancy Loss Unit we have since 2015, treated women with unexplained RPL with HCQ in off-label-use with a high compliance and showing a LBR of 70%^23^ compared to a LBR of 50 % for the placebo group in our latest db-RCT^24^. Based on the known association between maternal immunity and unexplained RPL, and our positive pilot off-label use of the HCQ for this group of patients, we have initiated a multicenter double-blind, randomized, placebo-controlled trial (db-RCT) to investigate the effect of HCQ treatment in women with unexplained RPL.

## STUDY OBJECTIVES

### Hypothesis

Treatment with HCQ can increase the LBR in women with unexplained RPL.

### Primary outcome

1. The difference in LBR (live birth defined as the birth of a child that survives the four-week neonatal period after birth) among women allocated to treatment with HCQ and those allocated to placebo treatment, without exclusion of any included patients (Intention to treat (ITT) analysis).

### Secondary outcome

1. The difference in LBR women allocated treatment with HCQ and those allocated placebo treatment, according to the protocol. PL due to chromosome abnormalities, ectopic pregnancy, neglect of treatment, induced abortion or those who withdraw from the treatment earlier than the protocol dictates do not fall under the category of treatment failure, and these are excluded in this secondary analysis (Per protocol (PP) analysis).
2. Birth weight, duration of pregnancy, Apgar-score five minutes after birth and the number of days admitted into a neonatal department. Patients with RPL have an increased risk of preterm birth, perinatal morbidities, and low birth weight^25,26^. Hence, it is relevant to investigate whether treatment with HCQ can reduce these complications, or if treatment with HCQ reduces the rate of PL at the expense of an increased rate of perinatal complications. The rate of perinatal complications is compared to the placebo group in this analysis.
3. Immune and inflammatory profiles and trajectories in women treated with HCQ vs placebo and women with live birth vs another PL. Pre-pregnancy inflammatory levels will be contrasted between women treated with HCQ and placebo to identify high responders. Furthermore, repeated measures analysis enables the understanding of pregnancy-complications and identification of high-risk groups at early stages, which may benefit from treatment.
4. Monitoring metabolic activity measured by ultra-low dose FDG-PET/CT scans in a sub-study. Metabolic activity correlates with grade of inflammation and will be measured in the uterus and immune-related tissue such as bone marrow, spleen, thymus, and lymph nodes.

## MATERIALS AND METHODS

### Study design

The study design is a randomized, double-blinded, placebo-controlled trial which compare HCQ treatment in women with unexplained RPL with minimum four consecutive PL or three PL including a second trimester PL to placebo treatment.

### Study setting

All patients referred to the RPL unit Capital Region (Rigshospitalet or Hvidovre Hospital) who according to inclusion and exclusion criteria are eligible are invited to participate in this db-RCT. Patient enrolment began in primo 2017 and so far, 149 out of 186 patients are included.

### Study participants and inform consent

Prior to the first visit, couples referred to the clinic will have completed a questionnaire, had a RPL blood panel taken and have seen an online information video introducing the trial. Based on the diagnostics at the clinic, eligible women diagnosed with unexplained RPL that fulfil the inclusion- and exclusion criteria with ≥4 consecutive PL or three PL including a second trimester PL, will be invited into the trial. If they are interested in the trial, they will be enrolled. In total four consent forms are signed, two by the woman, one by the partner and one which needs to be signed by both parents, allowing an extra examination of eyes and ears in case of a live birth. Additional consent from the woman is required for her to participate in the sub-study with ultra-low-dose PET/CT scans.

### Inclusion criteria

1. Unexplained RPL with ≥4 consecutive PL before 20 weeks of gestation or three PL including a second trimester PL (Pregnancy termination or confirmed ectopic localization of the pregnancy are not considered PL).

### Exclusion criteria

1. Age < 18 years or > 40 years.
2. Prominent uterine abnormalities detected by hysterosalpingography/hysteroscopy or hydrosonography.
3. Known prominent chromosome abnormalities for the couple trying to conceive.
4. A menstrual cycle < 23 days or > 35 days if the pregnancy is conceived naturally. If the woman received fertility treatment the menstrual cycle has no consequence.
5. Detection of positive lupus-anticoagulant or positive IgG/IgM for anticardiolipin-antibodies (≥10 GPL kU/l, measured at the same laboratories at the Capital Region of Denmark) or plasma homocysteine ≥25 microgram/l by repeated measurements 12 weeks apart before the pregnancy.
6. Positive HIV test or test indicating chronic hepatitis B or C.
7. Psoriasis, retinopathic or serious hearing deficiency (contraindication for treatment with HCQ).
8. Current chronic disease implicating a constant consumption of anti-inflammatory or potentially medicine harmful to the pregnancy/embryo.
9. Hgb ≤ 6.5 mmol/L, leucocytes < 3.5 E9/L, thrombocytes < 145 E9/L by the time of inclusion.
10. Previous HCQ treatment in relation to conceive.
11. > 1 previous live birth.
12. Previous participation in current study.

### Intervention

The intervention group are treated with 200 mg HCQ per day from minimum two months before pregnancy until 28 weeks of gestation or until PL, whichever comes first. Doses and duration are based on results from earlier studies showing it takes some time for treatment with HCQ to obtain maximum effect^27^. Furthermore, the half-life period for HCQ is 40-60 days^16^, which entails an effect of HCQ several weeks after withdrawal of the treatment. Doses are like the antirheumatics doses used on women pregnant in the first trimester^13^. The women randomized to the placebo-group are allocated 200 mg Lactose monohydrate per day.

### Allocation, randomization, and blinding

To ensure an equal distribution of included patients with four PL and ≥five PL in the intervention and placebo groups, the included patients will be allocated into two lists (randomized block design) at the Pharmacy of the Capital Region. The Pharmacy of the Capital Region dispenses and blinds the intervention and placebo treatment and stores the randomization code. The HQC and placebo treatment are both orally administered and will be packaged in identical gelatine capsules to ensure blinding. In addition, sealed envelopes with the treatment code for each patient included are prepared and stored in a locked cabinet in the clinic. If the decision to break the code for the patient is made, the investigator will inform the Pharmacy of the Capital Region that the concrete envelope has been broken.

### The treatment protocol (Fig 2)

Hgb, leukocytes and thrombocyte count are measured at the start of the treatment as well as approximately every six months. Included women must be examined by an ophthalmologist within three months from the treatment start, as well as after approximately one year of treatment. To optimize compliance, all patients are briefed prior to the initiation of treatment about the correct in-take of medicine, both orally and in writing, and side-effects as well as treatment compliance is monitored during hospital visits.

When an included patient report to be pregnant, the following intensive monitoring scheme is initiated (fig. 1):

**Figure 1:**
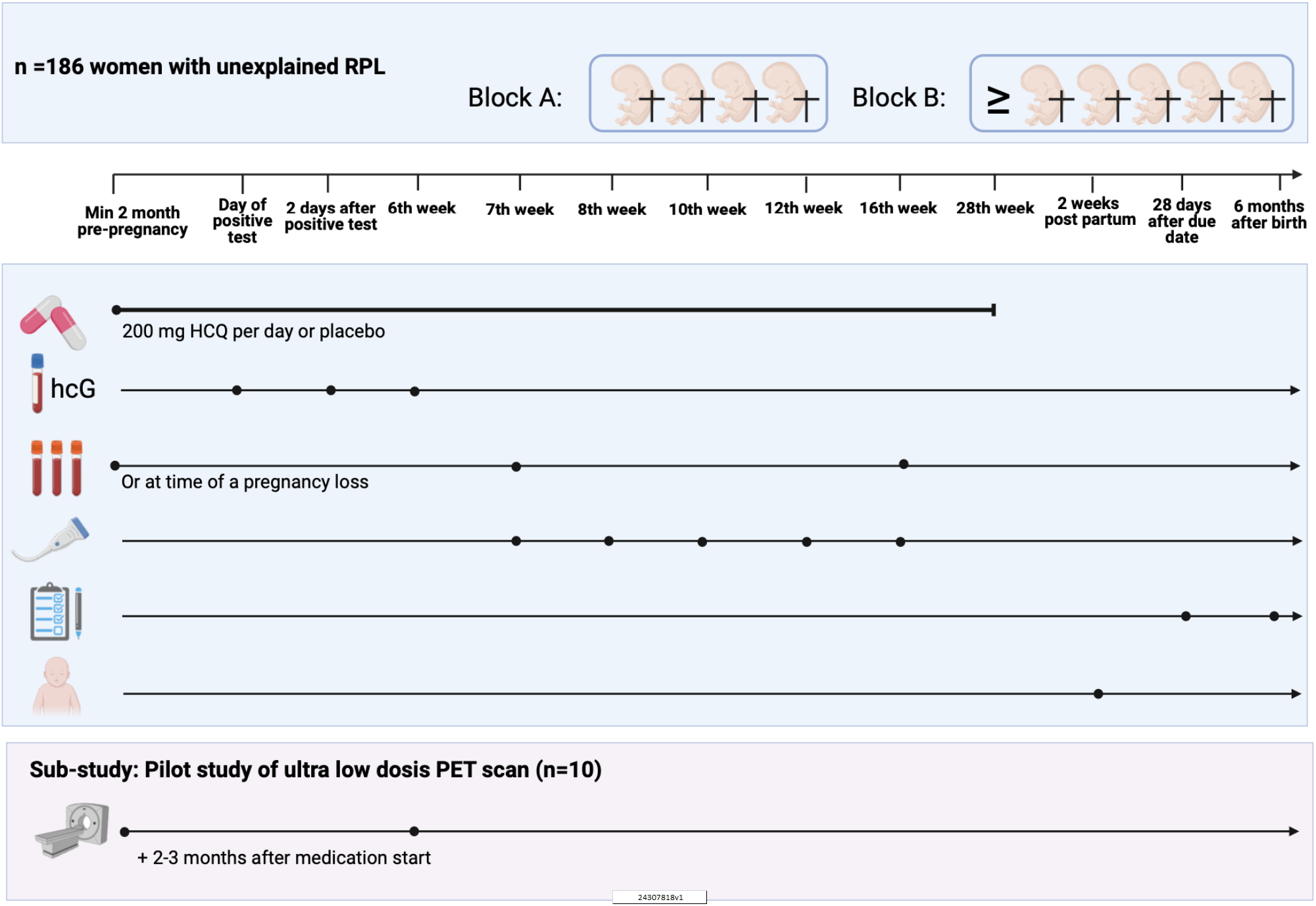
HCQ for unexplained RPL - a db-RCT study. Figure created in BioRender

◻ The day of positive pregnancy test: Blood samples examining human choriogonadotropin (hCG) and progesterone.
◻ 2 days after positive test (5th week of gestation): blood samples examining hCG.
◻ 6^th^ week of gestation: blood samples examining hCG
◻ 7^th^ week of gestation: first ultrasound scan
◻ 8^th^ week of gestation: second ultrasound scan
◻ 10^th^ week of gestation: third ultrasound scan
◻ 12^th^ week of gestation: fourth ultrasound scan
◻ 16th week of gestation: final ultra-sound scan at The Recurrent Pregnancy Loss Unit. Planning of follow-up examinations of the child at a local hospital (after birth) and extended screening of the baby’s ears and eyes during the first two weeks after birth for impairments of sight and hearing.

Additionally, all pregnant women in Denmark are offered a first-trimester scan to screen for trisomy 13, 18, and 21, and a second-trimester scan which, among other things, screens for fetal malformations. Information from these scans is collected through a database.

Questionnaire A: Trial participants who confirm an ongoing pregnancy by telephone-contact at 28 weeks of gestation, receive an electronic questionnaire by email 28 days after the due date. This contains questions regarding date and time for birth, method of birth, sex of the child, birth weight, pregnancy complications after 15 weeks of gestation, results of second-trimester scan, the baby’s state of health as well as the results of the extended screening of sight and hearing.

Questionnaire B: If questionnaire A confirms the birth of a child, questionnaire B will be sent to the trial participant when the child is six months old. This contains questions regarding the development of the child from birth up to six months.

#### Design for the sub-study with ultra-low dose PET/CT scans

Metabolic activity correlates with grade of inflammation and will be measured in the uterus, immune-related tissue such as bone marrow, spleen, thymus, and lymph nodes, and consequently we will detect signs of systemic inflammation using FDG-PET/CT before and during HCQ treatment, and in early pregnancy. Ten women from the db-RCT will be offered inclusion in this sub-study, at the time of inclusion in the db-RCT. FDG-PET/CT has the potential to detect low-grade inflammation in patients^28^. The ultra-sensitive Total Body PET scanner facilitates high quality PET-imaging with radiation doses of 1/10 of previous standards, and enables us to use FDG-PET/CT scans to visualize changes in the endometrium and immune tissue, and detect signs of systemic inflammation before and during pregnancy with and without HCQ treatment^29^

Included patients will have three FDG-PET/CT scans: 1) at inclusion, prior to starting the treatment, 2) after two-three months of treatment and, 3) after six weeks of gestation (fig. 1). Patients will be included until ten full sets of three PET/CT scans are obtained. Due to cyclic-conditional FDG-absorption in the uterus, the scan must take place late in the luteal phase or early in the follicular phase. The fasting participants (for minimum four hours) get an intravenous injection of 18F-FDG equivalent to 0.5 Mbq/kg one hour prior to the scan. For the third scan we use an AI-based attention correction during the scan instead of the CT, to keep the radiation dose to the fetus minimal.

### Withdrawal of participants or termination of the trial

If a patient becomes pregnant less than two months from initiating the study treatment or does not achieve pregnancy one year from the time of initiation of study medication, the patient is withdrawn from the trial, and will be replaced 1:1 by new patients included in the trial. In the case of side effects, adverse events (AE) or serios adverse events (SAE) related to treatment with HCQ, the investigator will evaluate whether to consent continuation in the trial or to open the sealed document with the randomization code for the patient. In case of SAE or persistent AE, the patients will be examined by doctors with relevant specialties.

Pregnant study participants excluded from the trial due to AE/SAE, experiencing PL or ectopic pregnancy are offered follow-up and continued care in the RPL unit.

The included patients will be excluded from the PP analysis if they are unable to continue the treatment until 12 full weeks of gestation (or minimum until four weeks before the gestation age of previous PL in the second trimester). Included patients who are uncontactable (repeatedly neglecting phone calls and messages in e-Boks) are excluded after three months with no contact. If the patient stops the treatment during the trial or if the patient is excluded, collection of the same information as in the questionnaires will be attempted. All data from excluded patients are included in the ITT-analysis.

### Research biobank

A research biobank is created in reference to the db-RCT and ends with the conclusion of the trial. Pending permission from the included patients in a separate consent form, surplus blood-sample materials will subsequently be transferred to a biobank for future research.

Five glasses of 6 ml blood (two glasses of plasma, two glasses of serum and one glass of full blood) will be collected up to four times per included patient for the research biobank. Samples will be taken once before the start of treatment (prior to pregnancy), twice during pregnancy and in case of a PL when that is diagnosed. Samples are collected, for later batch examination of immune and inflammatory profiles. Among others and given additional funding the following cytokines and immunological parameters will be examined: TGF-β, CSF1, GM-CSF, LIF, IL1, 4,5,6, 9, 10, 11,12 13, 15,17,23, TNF, IFN-γ, VEGF, FGF-a and b, hsCRP, PIF, D-vitamin, PTX3, true-immune culture analysis, complement Bb, C5A, tissue factor level + activity, complement analyses: classical pathway function, alternative pathway function, C1q, C4, C3, C3d, anti-C1q and T-/B-/NK cell analysis. Plasma samples from the first trimester and in case of a PL are biobanked for identification of circulating cell-free fetal DNA (cffDNA) for fetal genetic diagnosis in case of PL^2^.

### Data management

Ordinary procedures regarding quality control and quality assurance are pursued according to §3 and §4 in the GCP-proclamation. Data is collected in a single redcap database. The database is to be transferred to The National Archive no later than 2^nd^ of January 2042. We ensure all data is handled with confidence and consideration according to the GDPR rules.

### Safety

In general, HCQ is well tolerated and has few side effects. Nausea, vomiting, abdominal pain, headache, mood swings, dizziness, rash, itching, reduced appetite and rarely alopecia, tinnitus and cornea oedema is some of the known side effects^13^. Blood samples (Hgb, leucocytes and thrombocytes) will be taken before initiation of treatment and every six months. All included patients will be examined by an ophthalmologist within three months and after one year of treatment start. Since psoriasis, retinopathies, and severe hearing reduction is contraindications for HCQ treatment, they are exclusion criteria for participating in the db-RCT.

Reporting of side effects, AE and SAE will be performed according to guidelines form the Danish Medicines Agency. The patients will systematically be questioned about known and possible side effects, both at the first consultation, when the patient first discover pregnancy and at subsequent consultations (6 times until 15 weeks of gestation). In addition, the patient will have a direct phone number to the RPL Unit to ensure sufficient recoding of side effects besides planned consultations. The summary of Product Characteristic for HCQ will be used as reference in the assessment of side effects. All information concerning SAE will be recorded by the investigator to the Danish Medicines Agency and the National Committee on Health Research Ethics as soon as possible and at least seven days after their occurrence. No later than eight days after reporting a SAE, the investigator will brief all follow-up investigations concerning the SAE to the Danish Medicines Agency and the National Committee on Health Research Ethics. All recorded SAE will be accompanied by a comment on any consequences of the trial.

In the case of SAE attributable to HCQ, the investigator will decide if it is necessary to break the randomisation for the specific patient.

A proportion of AE’s as tiredness, nausea, abdominal pain, headache, mood swings, dizziness, itching, rash, and normal pregnancy related nausea will be registered in the patient’s journal, but not in the CRF. If the AE’s is of such magnitude that it affects normal daily activity the AE’s will be registered. Events related to the RPL diagnosis (vaginal bleeding, PL, intrauterine growth restriction, placental abruption, preterm birth, and preeclampsia) will not be considered as AE’s. Detection of a malformation in second or third trimester will be considered as a SAE. In case of an event that makes daily activity impossible, the patient will be monitored closely at the RPL clinic until the event has disappeared, or the monitoring passes to other departments, which will inform the investigator of the results of their evaluation.

#### Safety of the sub-study with ultra-low dose PET/CT scans

Participating women are exposed to a total radiation dose of 3-4 mSv (2-3 scans), equivalent to the annual background radiation in Denmark. The total risk of cumulative stochastic damage for the participating woman is 1:10.000^30^. In Denmark women have a 25 % risk of dying by cancer throughout their lifetime^30^. After participation in this study the estimated risk is 25,02%. Scanning during pregnancy exposes the fetus to radiation as well. The radiation dose for the fetus during injection of approximately 30 MBq is 21 microSv/MBq, equivalent to 0.6 mSv. Using AI-based attention correction during the scan instead of the CT, the radiation dose to the fetus is kept safely <1 mSV. The total risk of cumulative stochastic damage for the fetus is about 1:100.000. No risk for congenital malformation due to a radiation dose beneath 100 mSv has been documented

### Sample size and power calculation

The calculation of power is based on the LBR (50%) amongst the women in the placebo group, in our most recent db-RCT^24^ and an expected increase in LBR HCQ treated to 70% as seen in our pilot off label use^23^. The type I error (α) is estimated at 5 % and type II (β) error is estimated at 20 %. Consequently, 93 patients with unexplained RPL must be included in each group (intervention and placebo group) and 186 patients must be included based on the primary outcome measures.

#### The sub-study with ultra-low dose PET/CT scans

As the sub-study is clearly under-powered for the primary outcome in the db-RCT, the sub-study will provide an important feasibility evaluation and it will also be among the first time series of ultra-low dose PET/CT scans during pregnancy.

### Statistical analysis

In the ITT analysis the LBR (= number of pregnancies resulting in a live birth / total number of non-excluded pregnancies) in the intervention (HCQ) and placebo group is compared by X^^2^ test. A p-value < 0.05 is considered statistically significant. Relative risks with 95% CI for LBR in the intervention group vs placebo group will be calculated in both the ITT and PP analyses.

Loss-to-follow-up for the primary outcome, live birth, will be compensated through integration with the national birth registry. This will also allow us to extract other missing variables, such as birth weight etc.

Divergence to the original statistical plan will be reported. The trial concludes when all 186 fully evaluable patients are included in the trial and patients have given birth (after six months follow-up) or had a PL. The code will be broken after the statistical statement of the results is made, and subsequently the patients will be informed what treatment they were allocated. Interim analysis will not be executed.

The pre-defined inflammatory markers and immunological parameters will be analyzed to identify biomarkers for treatment response, using generalized linear models^31^. We will compare a model that includes only the treatment, with a model also including an interaction term with the potential biomarker and compare them using a likelihood ratio test. If the p-value for the LRT < 0.05, we conclude that the potential biomarker is predictive of response to treatment. Furthermore, it will be explored if an AI approach can successfully predict (treatment) success. We will utilize pre-trained transformer models that include healthcare utilization and socioeconomic parameters to predict treatment response^32^. Lastly, it will be investigated how inflammatory profiles differentiate a successful pregnancy from a PL, using unsupervised learning (e.g. principal component analysis or variational autoencoders).

#### Data analysis for the sub-study with ultra-low dose PET/CT scans

We will report simple descriptive results in relation to study medication (HQC or placebo), pregnancy, and pregnancy outcome.

## DISCUSSION / FUTURE PERSPECTIVES

Although PL and RPL are frequent, they remain poorly understood and it is time to accelerate action as recently concluded in a Lancet series documenting the alarming need for evidence-based treatments^26^. According to the European guideline for RPL (2022) recommended treatments are supported by low or very low-quality evidence, due to the lack of evidence-based investigations and treatments in RPL^33^. Currently there are very few documented treatments for patients with RPL. The current study will add quality data to the field as called for.

Until May 2024, three db-RCTs (including this) have been posted at clinical trial.gov including RPL women. The first was initiated by an American research group with inclusion of women with two or more PL. The trial was stopped as HCQ is considered a harmless drug and no patient agreed to the 50% risk of placebo treatment. The second was initiated by a French research group and included women with three or more PL. This study was stopped by the authorities due to two cases of polydactyly (without bone). The Danish Medicines Agency has reviewed the material and do not find reason to close this db-RCT.

By helping couples to achieve a healthy pregnancy and baby, the treatment will likely also help to reduce the high occurrence of stress and depression associated with RPL^34^. Furthermore, the insight into immunological mechanisms and inflammatory response associated with unexplained RPL before and during pregnancy, and related to immune treatment, will contribute to a more comprehensive understanding of the field, and aid future research in reproductive medicine and women’s health. The immune analyses can further elucidate the mechanisms behind the well described association of RPL and obstetrical complications in the following pregnancies^10^, and long-term adverse health outcomes such as autoimmune and cardiovascular diseases^35^. Finally, the trial can contribute to the understanding of HCQ’s anti-inflammatory and immune modulating characteristics beneficial to not only patients with RPL, but to greater use for all the immunological and inflammatory diseases where HCQ treatment is currently tested.

## PLAN FOR SHARING DATA AFTER THE CLINICAL TRIALS ENDS

Positive as well as negative and inconclusive findings will be published. Publication will be in accordance with laws regarding processing of personal data and The Danish Health Care Act (The Danish Health Care Act). When the clinical trial ends, the anonymized data will be made available to other researchers through public databases such as the Zenodo open data repository (CERN) or other equivalent databases. Trial data will be available for other researchers after they obtain data permission to do this.

## Data Availability

When the clinical trial ends, the anonymized data will be made available to other researchers through public databases such as the Zenodo open data repository (CERN) or other equivalent databases. Trial data will be available for other researchers after they obtain data permission to do this.

## ETHICS

The db-RCT including the PET/CT sub-study has been approved by The Regional Committee on Health Research Ethics (H-17001899), The Danish Medicines Agency (EudraCT-number 2016-004981-24), The Danish Data Protection Agency (RH-2017-236, I-Suite nr: 05693), and transferred to The Center for Data handling Capital Region of Denmark (p-2024-15960) in May 2024. The trial conforms to good clinical practice guidelines (GCP), is monitored by Jeanette Blom from The Danish GCP Units and has been registered at ClinicalTrials.gov (ClinicalTrials.gov ID NCT03305263). The trial complies with the trial protocol, ICH-GCP guidelines and present regulation from the authorities. We consider the trial to be fully compliant and in line with the Helsinki-declaration regarding bio-medical trials with human beings.

## REFERENCES

1. Guideline on the management of recurrent pregnancy loss. Published 2023. https://www.eshre.eu/Guidelines-and-Legal/Guidelines/Recurrent-pregnancy-loss

2. Schlaikjær Hartwig T, Ambye L, Gruhn JR, et al. Cell-free fetal DNA for genetic evaluation in Copenhagen Pregnancy Loss Study (COPL): a prospective cohort study. Lancet. 2023;401(10378):762–771. doi:10.1016/S0140-6736(22)02610-1

3. Ogasawara M, Aoki K, Okada S, Suzumori K. Embryonic karyotype of abortuses in relation to the number of previous miscarriages. Fertil Steril. 2000;73(2):300–304. doi:10.1016/S0015-0282(99)00495-1

4. Ticconi C, Pietropolli A, Di Simone N, Piccione E, Fazleabas A. Endometrial Immune Dysfunction in Recurrent Pregnancy Loss. Int J Mol Sci. 2019;20(21):5332. doi:10.3390/ijms20215332

5. Nielsen HS. Secondary recurrent miscarriage and H-Y immunity. Hum Reprod Update. 2011;17(4):558–574. doi:10.1093/humupd/dmr005

6. Christiansen OB, Nielsen HS, Kolte AM. Inflammation and miscarriage. Semin Fetal Neonatal Med. 2006;11(5):302–308. doi:10.1016/j.siny.2006.03.001

7. Christiansen OB, Nielsen HS, Lund M, Steffensen R, Varming K. Mannose-binding lectin-2 genotypes and recurrent late pregnancy losses. Hum Reprod. 2009;24(2):291–299. doi:10.1093/humrep/den377

8. Kolte AM, Steffensen R, Christiansen OB, Nielsen HS. Maternal HY-restricting HLA class II alleles are associated with poor long-term outcome in recurrent pregnancy loss after a boy. Am J Reprod Immunol. 2016;76(5):400–405. doi:10.1111/aji.12561

9. Nielsen HS, Witvliet MD, Steffensen R, et al. The presence of HLA-antibodies in recurrent miscarriage patients is associated with a reduced chance of a live birth. J Reprod Immunol. 2010;87(1-2):67–73. doi:10.1016/j.jri.2010.05.006

10. Nielsen HS, Steffensen R, Lund M, et al. Frequency and impact of obstetric complications prior and subsequent to unexplained secondary recurrent miscarriage. Hum Reprod. 2010;25(6):1543–1552. doi:10.1093/humrep/deq091

11. Fu B, Tian Z, Wei H. TH17 cells in human recurrent pregnancy loss and pre-eclampsia. Cell Mol Immunol. 2014;11(6):564–570. doi:10.1038/cmi.2014.54

12. Krieg S, Westphal L. Immune Function and Recurrent Pregnancy Loss. Semin Reprod Med. 2015;33(04):305–312. doi:10.1055/s-0035-1554917

13. Plaquenil (pro.medicin.dk). http://pro.medicin.dk/Medicin/Praeparater/622

14. Garner P, Gülmezoglu AM. Drugs for preventing malaria in pregnant women. In: Garner P, ed. Cochrane Database of Systematic Reviews. John Wiley & Sons, Ltd; 2006. doi:10.1002/14651858.CD000169.pub2

15. de Moreuil C, Alavi Z, Pasquier E. Hydroxychloroquine may be beneficial in preeclampsia and recurrent miscarriage. Br J Clin Pharmacol. 2020;86(1):39–49. doi:10.1111/bcp.14131

16. Rainsford KD, Parke AL, Clifford-Rashotte M, Kean WF. Therapy and pharmacological properties of hydroxychloroquine and chloroquine in treatment of systemic lupus erythematosus, rheumatoid arthritis and related diseases. Inflammopharmacology. 2015;23(5):231–269. doi:10.1007/s10787-015-0239-y

17. Gajić M, Schröder-Heurich B, Mayer-Pickel K. Deciphering the immunological interactions: targeting preeclampsia with Hydroxychloroquine’s biological mechanisms. Front Pharmacol. 2024;15. doi:10.3389/fphar.2024.1298928

18. Article “Drømmen om et barn: Marlene har været gravid 11 gange.” https://www.dr.dk/levnu/boern/droemmen-om-et-barn-marlene-har-vaeret-gravid-11-gange

19. Article “Baby born to woman who suffered 20 miscarriages.” Published online 2014. https://www.bbc.com/news/health-25775823

20. Lafyatis R, York M, Marshak-Rothstein A. Antimalarial agents: Closing the gate on toll-like receptors? Arthritis Rheum. 2006;54(10):3068–3070. doi:10.1002/art.22157

21. Weber SM, Levitz SM. Chloroquine Interferes with Lipopolysaccharide-Induced TNF-α Gene Expression by a Nonlysosomotropic Mechanism. J Immunol. 2000;165(3):1534–1540. doi:10.4049/jimmunol.165.3.1534

22. van den Borne BE, Dijkmans BA, de Rooij HH, le Cessie S VC. Chloroquine and hydroxychloroquine equally affect tumor necrosis factor-alpha, interleukin 6, and interferon-gamma production by peripheral blood mononuclear cells. J Rheumatol. 1997;24(1):55–60.

23. Eshre abstract book volume 31, supp 1 2016. Hum Reprod. Published online 2016. https://www.eshre.eu/Annual-Meeting/Helsinki-2016/Abstract-book

24. Christiansen O, Larsen E, Egerup P, Lunoee L, Egestad L, Nielsen H. Intravenous immunoglobulin treatment for secondary recurrent miscarriage: a randomised, double-blind, placebo-controlled trial. BJOG An Int J Obstet Gynaecol. 2015;122(4):500–508. doi:10.1111/1471-0528.13192

25. Wu CQ, Nichols K, Carwana M, Cormier N, Maratta C. Preterm birth after recurrent pregnancy loss: a systematic review and meta-analysis. Fertil Steril. 2022;117(4):811–819. doi:10.1016/j.fertnstert.2022.01.004

26. Quenby S, Gallos ID, Dhillon-Smith RK, et al. Miscarriage matters: the epidemiological, physical, psychological, and economic costs of early pregnancy loss. Lancet. 2021;397(10285):1658–1667. doi:10.1016/S0140-6736(21)00682-6

27. Promedicin.dk: Chloroquinderivater (inflammatoriske reumatiske sygdomme).

28. Hjuler KF, Gormsen LC, Vendelbo MH, Egeberg A, Nielsen J, Iversen L. Increased global arterial and subcutaneous adipose tissue inflammation in patients with moderate-to-severe psoriasis. Br J Dermatol. 2017;176(3):732–740. doi:10.1111/bjd.15149

29. Alberts I, Hünermund J-N, Prenosil G, et al. Clinical performance of long axial field of view PET/CT: a head-to-head intra-individual comparison of the Biograph Vision Quadra with the Biograph Vision PET/CT. Eur J Nucl Med Mol Imaging. 2021;48(8):2395–2404. doi:10.1007/s00259-021-05282-7

30. Retningslinjer om anvendelse af ioniserende stråling i sundhedsvidenskabelige forsøg (2011). https://nationaltcenterforetik.dk/Media/637858096811481954/Appendiks2.pdf

31. OlinkAnalyze: Facilitate Analysis of Proteomic Data from Olink. Published 2023. https://cran.r-project.org/package=OlinkAnalyze

32. Savcisens G, Eliassi-Rad T, Hansen LK, et al. Using sequences of life-events to predict human lives. Nat Comput Sci. 2023;4(1):43–56. doi:10.1038/s43588-023-00573-5

33. Bender Atik R, Christiansen OB, Elson J, et al. ESHRE guideline: recurrent pregnancy loss: an update in 2022. Hum Reprod Open. 2022;2023(1). doi:10.1093/hropen/hoad002

34. Kolte AM, Olsen LR, Mikkelsen EM, Christiansen OB, Nielsen HS. Depression and emotional stress is highly prevalent among women with recurrent pregnancy loss. Hum Reprod. 2015;30(4):777–782. doi:10.1093/humrep/dev014

35. Ranthe MF, Andersen EAW, Wohlfahrt J, Bundgaard H, Melbye M, Boyd HA. Pregnancy Loss and Later Risk of Atherosclerotic Disease. Circulation. 2013;127(17):1775–1782. doi:10.1161/CIRCULATIONAHA.112.000285

